# Impacts of teaching modality on U.S. COVID-19 spread in fall 2020 semester

**DOI:** 10.1101/2020.10.28.20221986

**Authors:** Syed Badruddoza, Modhurima Dey Amin

## Abstract

We study the impact of college reopening in Fall 2020 on county-level COVID-19 cases and deaths using the information of 1,076 randomly chosen four- and two-year undergraduate degree-granting colleges from the National Center for Education Statistics. These institutions include public, private nonprofit, and for profit schools from 50 US states and the District of Columbia. We match college and county characteristics using several methods and calculate the average treatment effects of three teaching modalities: in-person, online, and hybrid on COVID-19 outcomes up to two months after college reopening. In pairwise comparison, colleges reopened with in-person teaching mode were found to have about 35 percentage points more cases within 15 days of reopening, compared to those that reopened online, and the gap widens over time at a decreasing rate. Death rates follow the pattern with a time lag. However, colleges with hybrid mode reach up to the rates of in-person mode after some time. We also find that greater endowment and student population, bigger class size, and fewer Republican voters in the county are major predictors of choosing remote teaching modes over in-person.

## 1 Introduction

The COVID-19 pandemic and its containment measures have unprecedentedly affected human health and economic activities (e.g., McKibbin and Fernando 2020; Atkeson 2020). Countries across the world have implemented partial or full business and school closures to mitigate the spread of infection (Panovska-Griffiths et al. 2020; Di Domenico et al. 2020). Many U.S. colleges temporarily closed or switched to online in the spring semester, and over six out of ten colleges reopened in the fall semester with an in-person or a combination of in-person and online teaching plans (Gallagher and Palmer 2020; Picault 2021). College students mainly fall in the age cohort of 18 to 29 years, which has a lower death rate (0.4 percent) from COVID-19 (Wrighton and Lawrence 2020; Goodman et al. 2021), but a greater chance of socialization than other age cohorts. Although colleges have taken different measures–including symptom screening, contact tracing, cleaning and disinfecting, and mandatory mask policies–there is a chance that reopening colleges may spread the virus among faculty and staff serving students, as well as in the region around the college (Hubler and Hartocollis 2020).

Given the substantial risk of spread of infection from college campuses to the community, a policy question is, if and to what extent, switching to a more remote teaching modality helps in controlling communicable diseases. Although the severity of the pandemic somewhat reduced by the end of 2021, a study that analyzes the effect of different teaching modalities may offer directions in similar crises in the future—such as an expansion of a new variant of COVID-19. The current study applies a quasi-experimental approach to find the impacts of teaching modality on infection spread onto the neighboring areas. Intuitively, we match college and county characteristics to find a pair of similar colleges, where one chose in-person and the other online. Then we measure how choosing an in-person mode might have affected COVID-19 cases and deaths in the neighborhood as opposed to choosing an online mode. We perform similar exercises for two other pairs: in-person versus hybrid, and hybrid versus online. Results provide evidence that college reopening, especially within in-person teaching mode, increases COVID-19 cases and deaths over time.

Most studies so far have looked into the impact of elementary and secondary school closure, optimal reopening plans, and teaching strategy (e.g., Di Domenico et al. 2020; Panovska-Griffiths et al. 2020; Gandolfi 2020; Cohen et al. 2020). Walke et al. (2020) discuss the possible issues of U.S. college reopening and the prevention measures required on campuses. Some early studies find an aggregated negative effect of school closures on COVID cases and deaths in the community (e.g., Auger et al. 2020; Pan et al. 2020), whereas some others find little to no negative effect (e.g., UNICEF 2020; Herby et al. 2022). A more granular level study conducted by Chernozhukov et al. (2021) shows that in-person opening of K-12 schools relative to that with remote opening is associated with a 5 percentage point increase in the weekly growth rate of cases. However, remote teaching in colleges may relate to COVID-19 containment more than remote teaching in schools, because college students fall into age groups that are more vulnerable to the disease than their younger counterparts (Goodman et al. 2021). In a study close to ours, Andersen et al. (2020) use an event study under a difference-in-difference setting to investigate college openings’ association with cellphone-tracked human mobility and COVID-19 cases in the counties of the campuses. Their preliminary results suggest that, reopening increased county COVID-19 cases by 1.7 daily per 100,000 county residents, and in-person teaching created greater visits and higher cases during the reopening weeks, but no contribution from online teaching was found. Nevertheless, most of these studies do not consider the fact that the choice of reopening and teaching modality is endogenous to observed cases. One of the assumptions of the difference-in-difference method is that the allocation of intervention was not determined by outcome, which is violated in this case because teaching modalities were often chosen by colleges based on pre-existing COVID rates in the region. Moreover, COVID cases can be influenced by required testings during the reopening weeks, and deaths may take longer than two weeks to appear in the data. Additionally, there can be other institutions (e.g., K-12 schools) affecting both COVID outcomes and the decision of instructional modalities. Our primary source of identification is that different colleges reopened on different dates, allowing for a clear comparison. We utilize data from various sources, including cellphone mobility data for comprehensive matching; and we use methods that account for multidimensional differences among colleges and re-weight the sample based on the college’s possibility of selecting an instructional modality.

## 2 Method

We are interested in finding the effects of teaching modality on two COVID-19 outcomes: (1) new COVID-19 cases reported in the county, and (2) new COVID-related deaths in the county. The impact of teaching modality is assessed on county-level outcomes (and not on the college level) because students may spread the virus to non-students and immunocompromised people in the community, which is a critical policy concern. First, we specify a simple relationship between college reopening in the fall semester and the outcome as below,

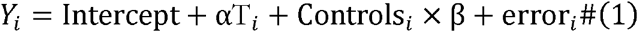

where, *Y*_*i*_ is a COVID-19 outcome, *i =* 1, …, *N* indicate colleges and *T*_*i*_ is a binary treatment variable that represents teaching modality. We use three specifications of *T*_*i*_ separately: (1) in-person=1, online=0, (2) in-person=1, hybrid=0, and (3) hybrid=1, online=0 for pairwise comparison of teaching modalities. That is, for each of three regressions we only include the observations with respective teaching modalities. The vector of control variables includes observed college- and county-level covariates such as college endowments or percent of the population who stayed at home, and *β* is a vector of respective parameters. Our parameter of interest is *α*, which shows the impact of a mode of instruction on an outcome variable, e.g., whether switching from online to in-person has an impact on the COVID-19 outcomes. The equation above would have been identified if colleges were homogeneous and were randomly assigned a teaching mode (Barnow et al. 1981). In practice, the treatment assignment is not random, because colleges choose the mode of teaching based on their distance education capacities, observed COVID-19 cases, and many other characteristics. The probability of adopting a teaching modality may vary considerably across colleges. Intuitively, if we could select two colleges with identical institutional and regional features, except one went in-person and the other online, we could measure the difference in COVID outcomes induced by the teaching modality.

There are some other estimation issues. First, many areas had high COVID-19 cases in the spring and summer semesters. Due to the exponential nature of the disease spread, places with more initial cases will experience faster growth. Moreover, teaching modality in the fall semester may depend on existing cases, thus generating a reverse causality. Second, there can be observed and unobserved college and county features that affect both teaching modality and COVID-19 outcomes, creating an omitted variable bias. Third, a measurement error may occur because infections are reported less during the weekend and more on weekdays, and the incubation or survival period varies. In addition, colleges reopening with in-person teaching elements may require mandatory COVID testing, which leads to an increase in the number of cases. Finally, a county may have multiple colleges or K-12 schools that determine the presence of people in the region, which in turn may affect both college teaching modality and COVID spread. We take several measures to address these problems. For instance, we control for cellphone mobility, many observed college and county characteristics, and use percentage changes of the outcome variables to eliminate college and county-level unobserved temporal fixed effects. We also check the outcomes for up to two months and control for the presence of other colleges in a county. Attempts to address other estimation issues are described below^1^.

One source of identification is that different colleges reopened on different dates between July and October of 2020. Therefore, we can isolate the effect of reopening on COVID-19 from the aggregate trend. A bigger identification problem is the presence of other colleges, K-12 schools, clubs, churches, etc. in the county that are unobserved in the data. Trips to these areas may affect the disease spread, as well as the selection of college teaching modalities. Thus, omitting the level of crowdedness or trips made in the county will lead to omitted variable bias. We follow Chernozhukov et al. (2021) to address the problem. They used foot traffic in K-12 schools captured by cellphone mobility data to measure if the school is operating in-person (more foot traffic) or online (less foot traffic). Notice that, even if we control for the foot traffic in K-12 schools, we still need to control for the foot traffic in numerous other places to avoid the omitted variable bias, i.e., we must add many other variables in the regression model. A parsimonious way to address the issue is to control for the percentage of the county population who stayed home, recorded by their cellphone locations, according to the following logic:

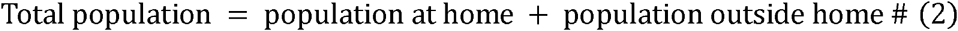

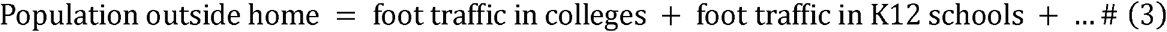

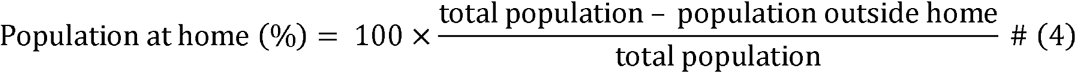

where, being outside is specified by staying outside the home for longer than ten minutes^2^. Thus, controlling for the percentage of the population who stayed home benefits the estimation process in many ways, because it comprehensively covers people who are running errands, going for a COVID test, going to schools, colleges, and any other places. We give two examples below.

First, if there are multiple colleges in the county and we do not observe whether the unobserved colleges are in-person or online, cellphone mobility captures the variation. If the unobserved college reopens in-person, average cellphone mobility in the county should increase more than if it reopens online. On the flip side, the percentage of the population staying home will decrease (eq. 4). Then the increase in COVID outcomes will be explained by the percentage of the population staying outside in various places, and will not corrupt the coefficient with the teaching modality for the observed college. Moreover, if the college teaches in-person but follows a strict “closed” campus policy, cellphone mobility captures that as well. Assume two colleges both teaching online, but one has more students returned to campus for an open campus policy. The cellphone mobility controls for the variation in COVID outcomes unexplained by teaching modality.

Second, Chernozukov et al. (2021) found a 0.09 correlation between K-12 school visits and college visits during the reopening weeks. Thus, if K-12 school reopening is correlated with college teaching modality, our estimation will be biased. Following Chernozukov et al. (2021), the reopening of K-12 schools will reflect in their foot traffic, and controlling for cellphone mobility will capture the unobserved variation in COVID outcomes, which allows us to estimate the effect of college teaching modality. Infection rates in neighboring counties will affect a county via population mobility, so a splillover effect will be controlled for as well.

What does the coefficient with the teaching modality variable capture? Assume two comparable colleges in two counties have the same foot traffic, and counties are similar in all other aspects related to COVID and college teaching modalities, both of the colleges have students on campus and similar average cellphone mobility per day, but one of them reopened online and the other in-person. Our estimate gives the difference between covid cases in two counties generated by the teaching modalities only: the effect of students attending an in-person class as opposed to an online one, holding everything else fixed.

To deal with the heterogeneous starting values of COVID, measurement errors (e.g., more testing on weekdays), and threshold selection issues (e.g., incubation period), we use two-week intervals: 0-15 days after reopening, 15-30 days after, 30-45 days after, and 45-60 days after reopening; and run separate cross-section regressions for each subsample. The two-week period is selected based on the incubation periods (e.g., Paul and Lorin 2021). The benchmark of percentage calculation is 0-15 days before reopening; for example,

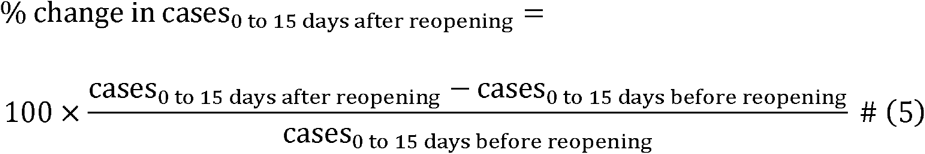

and so on. We chose two weeks intervals, because doing so gives COVID-19 enough time to appear in the outcomes, and we did not go beyond 60 days because the effects may be contaminated by other exogenous factors.

More testing will result in more cases. The 15-days interval approach, combined with the cellphone mobility above, helps us isolate the effect of increased COVID testing during reopening weeks in colleges with in-person modes. This is intuitive because, (1) testing requirements for college reopening are less likely to go beyond the first month after the semester starts, so a statistically significant *α* beyond the first 30 days is more likely to represent the contribution of teaching modality, (2) cellphone mobility reflects trips to the testing center as well, hence helps to identify *α*, and (3) COVID-related deaths are not affected by testing requirements, and incubation-to-death period may take longer than a month. So a positive significant *α* for COVID-related deaths after 30 days presents a more convincing case.

The next challenge is to ensure the comparability between colleges. We implement various matching methods described below to address this problem. We employ five methods and check if the results are robust: (1) bivariate Ordinary Least Squares (OLS) where *β* = 0 in the model above, (2) multivariate OLS where *β* ≠ 0, (3) propensity score matching (PSM), (4) Nearest Neighbor matching (NN), (5) and Kernel Multivariate Distance Matching (KMDM). The first two methods are conventional. The latter three methods focus on finding “statistical twins” for each observation in the treatment group from the control group with similar values of the covariates. The average treatment effects can be calculated as the mean of differences between the observed values in the treatment group and the imputed counterfactual values. We present a non-technical version of the matching methods below.

Rosenbaum and Rubin (1983, 1984) discuss propensity score matching (PSM) to adjust the probabilities for the differences in pre-treatment variables. A propensity score is the conditional probability of receiving the treatment given the pretreatment variables, i.e., *P*(*T*_*i*_ = 1|*X*_*i*_). The probabilities of treatments generated are used to create weights that adjust the pre-treatment imbalances. For example, if a college has a high probability of choosing online than in-person, greater weights are assigned to in-person than online during the estimation. PSM is usually calculated using logistic regression. For a recent technical discussion, see Imbens (2000) and McCaffrey et al. (2013).

Although finding a match with PSM is relatively easier, many studies argue that matching with propensity scores can be misleading as it ignores the multidimensional differences between two observations and simplifies them into one dimension—the score (e.g., King and Nielsen 2019). The nearest neighbor (NN) matching adopts a more multidimensional approach and uses some (default being one) closest observations in the control group. In the case of ties, NN uses all ties as matches. Conversely, a single control observation can be used many times with replacement. Matching with more than one continuous covariate may induce bias, so we apply a bias-adjusted NN matching proposed by Abadie and Imbens (2011).

Our final matching technique, KMDM, is similar to NN, but uses a certain bandwidth for multivariate matching instead of comparing with the closest neighbor. Simulation studies show that KMDM tends to outperform PSM because it approximates fully blocked randomization that is relatively more efficient (King and Nielsen 2019). We apply KMDM where observations in treatment and control groups are matched using Mahalanobis distance, and use the Epanechnikov kernel function to assign larger weights to controls with smaller distances. Following Huber et al. (2015), we choose 1.5 times the 90 percent quantile of the non-zero distances in finding a pair. The use of KMDM is more appropriate in our case, given the multidimensional heterogeneity among colleges and counties. See Jann (2017) for further discussion on the advantages of KMDM over other methods.

## 3 Data and descriptive statistics

For college characteristics, we utilize the Integrated Postsecondary Education Data System (IPEDS) survey annually released by the National Center for Education Statistics (NCES 2020). The IPEDS survey includes information on four- and two-year undergraduate degree-granting colleges and universities from all 50 U.S. states and the District of Columbia. Variables include tuition and fees, enrollment, number of degrees and certificates conferred, number of employees, financial statistics, and so on. Over six thousand such Title IV institutions (i.e., institutions that process U.S. federal student aid) were listed by the NCES in 2020. These institutions include public, private nonprofit, and private for profit schools that provide postsecondary education or training beyond the high school level. We aimed at selecting a thousand colleges for our analysis and randomly chose 1,100 of them using a random number generator under uniform distribution. The extra 100 colleges were selected to ensure that dropping missing values did not decrease the sample size below 1,000. Merging this dataset with other datasets, as described below, eventually gives 1,076 colleges after dropping the missing observations. Among the selected colleges, 41 percent are public, 42 percent are private non-profit, and the remaining 17 percent are private for profit.

Data on teaching modality and the official start date of the fall 2020 semester were manually collected between July and December of 2020 by authors from respective college websites.^3^ Over 90 percent of the colleges mentioned their latest teaching modality between June and September 2020. Information for the remaining colleges was obtained from contemporary campus news or local news. Some examples of search phrases we used are, “XXX university fall 2020 covid information reopening plan new” or “XXX college restarts in-person class in fall 2020” or “XXX institute fall 2020 plan president announcement updated”. We categorize fall reopening plans into three types: (1) in-person, (2) online, and (3) hybrid. Colleges in the “in-person” group started the fall 2020 semester with face-to-face classes and open residence halls, and may include few online delivery materials; colleges in the “online” group primarily offered online classes with some exceptions for lab components and may have some students on campus; and colleges in the “hybrid” group either divided the class into online and in-person section, or switched the teaching mode on a rolling basis, or offered courses with both in-person and online access. Thus, colleges that changed teaching modalities during the sampled period were considered hybrids. We understand this might create noise in the hybrid category, but we kept the in-person and online categories pure (either full in-person or fully online), as their difference is the main interest of this research. Moreover, the definition of hybrid substantially varies from one college to another, so it is more convenient to utilize in-person and online modes to infer the effects of hybrid modes than to study all types of hybrid modes. In our sample, 406 colleges (37.73 percent) taught online, 386 (35.87 percent) taught in person, and the remaining 284 (26.39 percent) followed a hybrid method.

We obtain daily new COVID-19 cases and deaths at the county level from the New York Times (NYTimes 2020) to stay consistent and comparable with similar studies conducted before (e.g., Fox et al. 2020; Andersen et al. 2020; Chernozhukov et al. 2021). Moreover, the NY Times data and CDC data are highly correlated (Chernozhukov et al. 2021). Figure 1 simply plots the changes in COVID-19 cases and deaths as a percent of 15 pre-semester days grouped by college teaching modality. We took the percentage changes to remove the influence of initial values, and to get rid of the college and county-level idiosyncratic features. Different colleges started the fall 2020 semester on different dates from the end of July to October. The earliest reopening date for the fall semester is July 20^th^ in our sample whereas the latest date is October 24^th^. Figure 1 matches the reopening dates, and suggests that COVID-19 cases were exponentially growing in the fall semester regardless of teaching modality, and COVID-related deaths followed the pattern. However, colleges teaching online are located in counties that had slower growth of cases after the college reopening date, on average. Cases were slowly growing for colleges with hybrid teaching modes as opposed to colleges with in-person modes. After 30 days, the pattern of cases is reflected in the pattern of deaths. On average, counties where colleges taught in hybrid or in-person mode had greater COVID-related deaths than counties where colleges taught online. However, an econometric analysis is required to find the impact of teaching modality on COVID outcomes.

**Figure 1.**
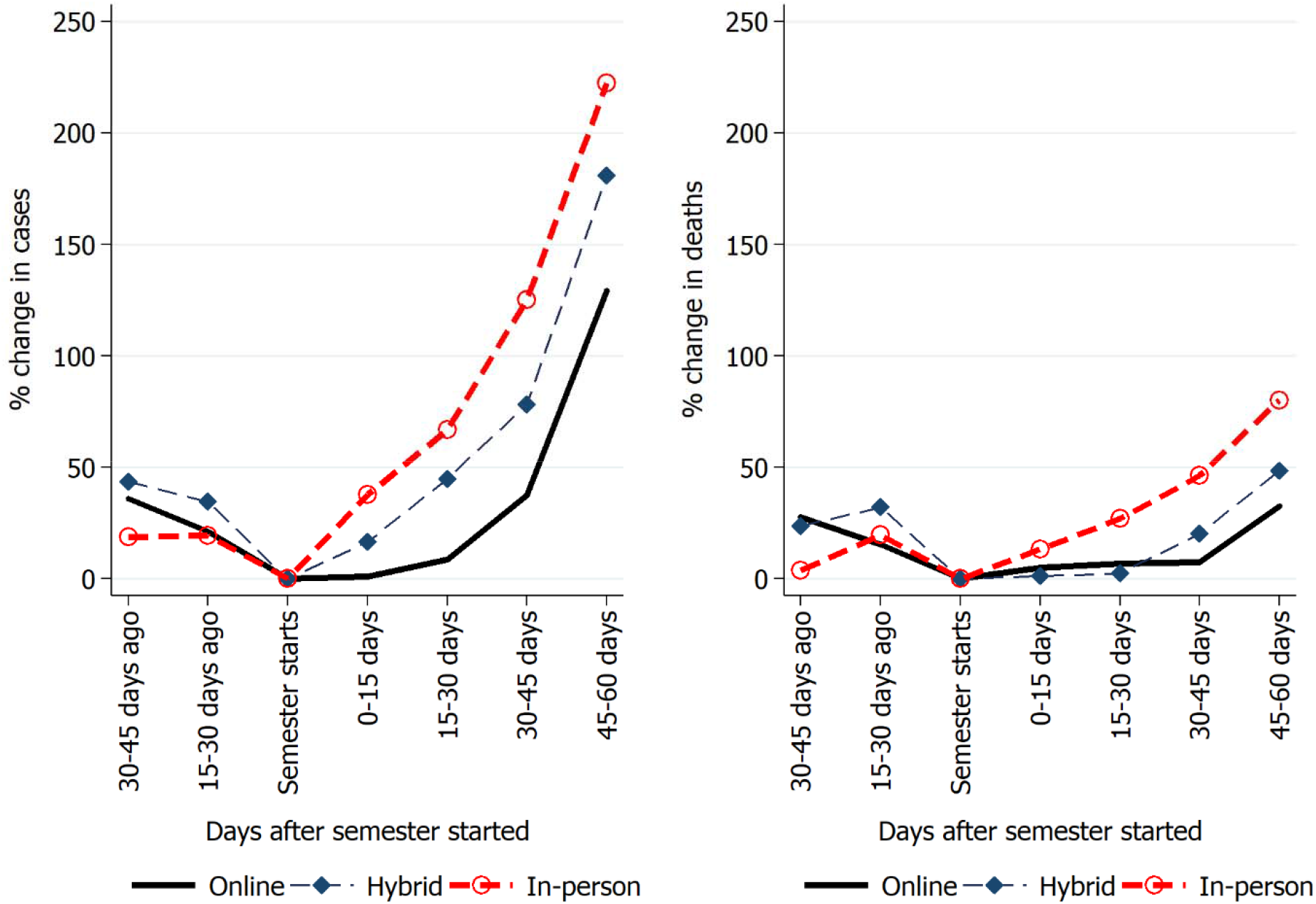
Growth of COVID-19 cases by college teaching modality in fall 2020 semester. Note: Figures are averages across colleges. Daily data of 1,076 colleges from July to October 2020. See the data section for details. Source: COVID outcomes are obtained from the New York Times (2020), and teaching modalities were collected by authors from the respective college websites.

We select several control variables for the econometric analysis based on the literature. Table 1 provides variable descriptions and sources, and Table 2 presents summary statistics of variables. As discussed in the method section, one of the most important control variables is the average percentage of the county population that stayed home. We collect this variable from the Bureau of Transportation Statistics (BTS 2020). The variable records the seven-day moving average of cellphone mobility at the county level, such that “staying home” is defined by cellphones that do not go outside the home for longer than 10 minutes. Intuitively, we get a percentage of county population that are not home for each ten minutes, then average the figures to get one number for a day, then calculate the seven-day moving averages. As shown in Table 2, fewer percentages of the county population (24.21 percent) stayed home under the in-person category, compared to hybrid (25.05 percent) and online (26.41 percent). The similar staying home rates across teaching modalities makes the covariate balancing more appropriate. Omitting the stay-home variable makes the effects of a teaching mode on COVID-19 outcomes biased, because COVID-19 outcomes may increase if fewer people stay home, given that cellphone mobility is associated with teaching modalities in schools (Chernozhukov et al. 2021).

**Table 1.**
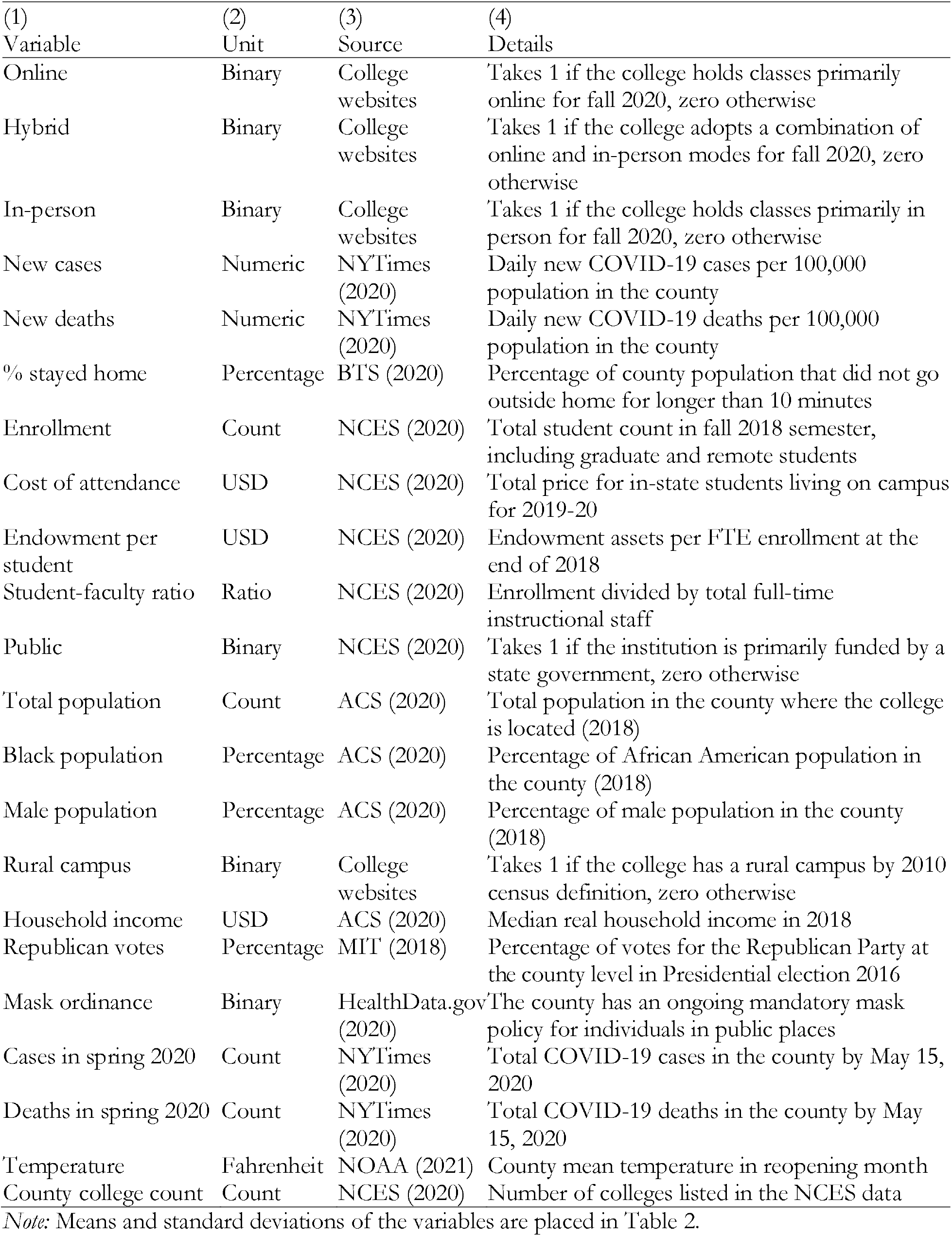
Variable description.

**Table 2.** Summary statistics.

| (1)<br>Variable | (2)<br>Online | (3)<br>Hybrid | (4)<br>In-person | (5)<br>Overall |
| --- | --- | --- | --- | --- |
| Daily new cases | 247.00<br>(558.23) | 100.04<br>(282.69) | 72.43<br>(248.92) | 145.59<br>(408.97) |
| Daily new deaths | 4.12<br>(10.54) | 1.62<br>(5.98) | 1.09<br>(4.39) | 2.38<br>(7.75) |
| Online |  |  |  | 0.38<br>(0.48) |
| Hybrid |  |  |  | 0.26<br>(0.44) |
| In-person |  |  |  | 0.36<br>(0.48) |
| % stayed home | 26.41<br>(4.06) | 25.05<br>(4.33) | 24.21<br>(4.00) | 25.27<br>(4.22) |
| Enrollment | 8,828.61<br>(9,991.36) | 6,307.21<br>(8,272.99) | 4,417.95<br>(6,972.39) | 6,580.85<br>(8,754.75) |
| Cost of attendance | 44,980.83<br>(21,312.56) | 46,178.28<br>(19,431.71) | 41,089.31<br>(16,628.42) | 43,757.31<br>(19,059.39) |
| Endowment per student | 61,279.10<br>(347,471.23) | 28,545.41<br>(62,887.43) | 59,892.61<br>(367,714.37) | 51,811.22<br>(307,016.55) |
| Student-faculty ratio | 41.71<br>(30.23) | 28.11<br>(22.71) | 29.18<br>(25.53) | 33.61<br>(27.44) |
| Public | 0.60<br>(0.49) | 0.38<br>(0.49) | 0.33<br>(0.47) | 0.44<br>(0.50) |
| Total population | 1,578,217<br>(2,620,932) | 616,050<br>(973,588) | 444,525<br>(1,028,931) | 917,566<br>(1,867,046) |
| Black population | 14.57<br>(15.77) | 13.68<br>(14.44) | 11.89<br>(13.14) | 13.37<br>(14.55) |
| Male population | 49.13<br>(1.28) | 49.13<br>(1.19) | 49.14<br>(1.27) | 49.14<br>(1.26) |
| Household income | 64,587.54<br>(17,710.66) | 59,805.99<br>(15,156.93) | 56,796.75<br>(14,395.72) | 60,526.88<br>(16,253.30) |
| Republican votes | 38.94<br>(16.72) | 46.09<br>(16.25) | 52.97<br>(14.53) | 45.86<br>(16.93) |
| Mask ordinance | 0.86<br>(0.35) | 0.80<br>(0.40) | 0.71<br>(0.45) | 0.79<br>(0.41) |
| Rural campus | 0.16<br>(0.37) | 0.22<br>(0.41) | 0.35<br>(0.48) | 0.24<br>(0.43) |
| Cases in spring 2020 | 12,706.16<br>(23,007.05) | 5,356.90<br>(10,396.52) | 3,576.15<br>(10,177.45) | 7,498.42<br>(16,800.38) |
| Deaths in spring 2020 | 594.02<br>(1,038.35) | 288.29<br>(568.51) | 70.89<br>(5.298) | 370.25<br>(789.65) |
| Temperature (F) in reopening month | 72.37<br>(6.14) | 71.38<br>(6.07) | 70.89<br>(5.30) | 71.58<br>(5.86) |
| County college count | 6.697<br>(8.462) | 3.637<br>(3.579) | 3.078<br>(3.748) | 4.591<br>(6.174) |
| Observations (Colleges) | 406 | 284 | 386 | 1,076 |

*Note:* Figures show averages across colleges. Standard deviations are placed in parentheses. Table 1 provides variable descriptions.

We include five college-level controls: (1) enrollment, (2) cost of attendance, (3) endowment per student, (4) student-faculty ratio, and (5) public versus private dummy. The total enrollment variable controls for the size of the college, which can be an important determinant of disease spread (e.g., Fox et al. 2020). Endowment per student and cost of attendance respectively capture the financial capacity of the college and the average affluence of its students, both are critical determinants of technology adoption in schools (e.g., Brahmasrene and Lee 2012; Leu et al. 2015; Sun and Chen 2016; Gallagher and Palmer, 2020). The variable on student-faculty ratio is an indirect measure of the average class size (De Paola et al. 2013), that may affect both case spread and remote teaching decisions. We also include a binary variable on whether the college is public or not because public colleges can be subject to COVID containment rules and regulations. Moreover, many public schools may have administrative or financial constraints to abruptly change teaching modes. Table 2 indicates that these college characteristics vary across teaching modalities.

Control variables on county characteristics were extracted from the American Community Survey (ACS 2020). One of the most important variables is the total population, which primarily determines the rate of disease spread (Siedner et al. 2020). We further added the percentage of Black population and the percentage of men in the county to capture their disproportionately higher COVID rates (e.g., Millett et al. 2020; Kim et al. 2021). Real household income controls for the average financial capacity of the county population, including students, to adopt new technologies and deal with income shocks during COVID (e.g., Tan et al. 2021). The binary variable on rural campus is added following the definition of US census because the rural location may affect the capacity and decisions regarding teaching modality. A variable on the existing mask ordinance was also included because mandatory mask mandates were found to have a negative association with case growth (Chernozhukov et al. 2021). Individual mask policies in county and state were obtained from HealthData.gov (2020). For some states, decisions regarding public university reopening were determined by central agencies, so we include state fixed effects in all models.

We also added information on county-level shares of Republican votes in Presidential Election 2016 from the MIT Election Data and Science Lab (MIT 2018) to take residents’ perception of COVID-19 risk into account (e.g., Tyson 2020). Two other critical county-specific variables are COVID-19 cases and deaths in the spring 2020 semester, because it was the last regular semester that gave the college administration a signal about the disease situation in the community. Higher cases and deaths in spring while the campus was open may influence college administrations to switch to online teaching modes (Gallagher and Palmer 2020). Some studies find an association between temperature and COVID-19 spread (e.g., Livadiotis 2020), so the average temperature in the county during the reopening month was included in the model from the National Oceanic and Atmospheric Administration (NOAA 2021) data. Finally, the total number of colleges in the respective county (including the sampled college) was included in the regression from NCES data, in order to remove the spillover effects, i.e., the contribution of other colleges to COVID outcomes. We admit that the data may not include all offices and institutions that may affect both COVID cases and teaching modality, but controlling for cellphone mobility together with college count and other identification strategies above should isolate the influence of relevant institutes from our analysis.

A required condition for estimating treatment effects is distributional similarities of pre-treatment covariates across the treatment groups (McCaffrey et al. 2013). Table 2 indicates considerable variation across teaching groups. The difference justifies the use of matching in our analysis. Tests for the distributional equality of matching variables between the treatment and control groups are placed in the appendix (see Figure A1). For PSM, these control variables are used to find the determinants of teaching modality using logistic regressions. The matching process should only contain variables that are measured at baseline, because variables measured at around the treatment may be influenced or modified by treatment (Austin 2011). Therefore, we selected the determinants from the latest academic year available before the fall 2020 semester.

## 4 Results

Tables 3 and 4 respectively show the average treatment effects of teaching modalities on COVID-19 cases and deaths. We begin with simple bivariate OLS to test the correlation, then control for other covariates, and then use three matching models. Apart from the bivariate OLS (Model 1), all control variables were used in all estimates (Models 2-5).

**Table 3.** Average treatment effects on COVID-19 cases. (Dependent variable: % change in cases compared to cases in two weeks before reopening date)

| Model | Treatment | (1) | (2) | (3) | (4) |
| --- | --- | --- | --- | --- | --- |
|  |  | 0-15 Days | 15-30 Days | 30-45 Days | 45-60 Days |
| 1. Biv. OLS | 1. In-person=1 online=0 | 18.291**<br>(9.259) | 22.265<br>(13.625) | 38.044<br>(24.626) | 24.598<br>(29.549) |
|  | 2. In-person=1 hybrid=0 | 12.768<br>(10.273) | 3.537<br>(20.489) | 27.221<br>(30.055) | 21.753<br>(34.139) |
|  | 3. Hybrid=1 online=0 | 5.552<br>(8.497) | 18.532<br>(16.705) | 22.661<br>(17.007) | 19.474<br>(23.512) |
| 2. Mult. OLS | 1. In-person=1 online=0 | 33.152**<br>(15.203) | 39.162*<br>(21.994) | 60.235<br>(37.523) | 4.856<br>(38.79) |
|  | 2. In-person=1 hybrid=0 | 23.629*<br>(12.96) | 18.091<br>(21.659) | 39.057<br>(33.033) | 3.023<br>(37.159) |
|  | 3. Hybrid=1 online=0 | 4.261<br>(12.189) | 10.94<br>(18.592) | 20.976<br>(16.925) | 39.665<br>(30.561) |
| 3. PSM | 1. In-person=1 online=0 | 35.052***<br>(13.174) | 36.061*<br>(19.77) | 49.915<br>(33.179) | 21.246<br>(33.564) |
|  | 2. In-person=1 hybrid=0 | 22.983*<br>(13.02) | 17.79<br>(22.032) | 41.661<br>(33.546) | 15.955<br>(39.944) |
|  | 3. Hybrid=1 online=0 | 6.607<br>(10.74) | 3.782<br>(16.098) | 18.706<br>(19.398) | 26.166<br>(44.361) |
| 4. NN | 1. In-person=1 online=0 | 24.459*<br>(12.881) | 26.044*<br>(14.811) | 46.144*<br>(25.293) | 48.135*<br>(27.656) |
|  | 2. In-person=1 hybrid=0 | 20.004**<br>(9.765) | 18.501<br>(14.026) | 41.576**<br>(20.643) | 48.599*<br>(28.815) |
|  | 3. Hybrid=1 online=0 | 7.311<br>(11.304) | 11.901<br>(18.141) | 10.299<br>(13.583) | 15.661<br>(21.643) |
| 5. KMDM | 1. In-person=1 online=0 | 35.014***<br>(10.631) | 54.036***<br>(15.375) | 79.443***<br>(28.319) | 81.676**<br>(36.443) |
|  | 2. In-person=1 hybrid=0 | 18.265<br>(12.887) | 12.657<br>(22.913) | 35.801<br>(31.083) | 21.845<br>(40.897) |
|  | 3. Hybrid=1 online=0 | 14.813<br>(10.944) | 27.690<br>(19.227) | 33.37**<br>(16.514) | 53.353*<br>(28.153) |

*Note:* Robust standard errors in parentheses. \*\*\* $p < 0.01$ , \*\* $p < 0.05$ , \* $p < 0.1$ . N=1,076. Model 1 has no control variable. Models 2-5 have the same control variables. Control variables are listed in Table 5 and matching performance is reported in the appendix (Figure A1). State fixed effects were also used but not reported. Biv.=Bivariate, OLS=Ordinary Least Squares, Mult.=Multiple, PSM=Propensity Score Matching, NN=Nearest Neighbor, KMDM=Kernel Multivariate Distance Matching.

**Table 4.** Average treatment effects on COVID-19 deaths. (Dependent variable: % change in deaths compared to deaths in two weeks before reopening date)

| Model | Treatment | (1) | (2) | (3) | (4) |
| --- | --- | --- | --- | --- | --- |
|  |  | 0-15 Days | 15-30 Days | 30-45 Days | 45-60 Days |
| 1. Biv. OLS | 1. In-person=1 online=0 | 6.957<br>(8.163) | 10.057<br>(10.674) | 10.966<br>(11.535) | 1.841<br>(15.012) |
|  | 2. In-person=1 hybrid=0 | 14.127<br>(8.809) | 22.991**<br>(11.044) | 13.781<br>(12.559) | 17.915<br>(17.359) |
|  | 3. Hybrid=1 online=0 | 3.847<br>(7.045) | 0.669<br>(9.995) | 9.568<br>(10.358) | -1.631<br>(13.33) |
| 2. Mult. OLS | 1. In-person=1 online=0 | 13.789<br>(11.488) | 23.266*<br>(13.931) | 6.053<br>(14.993) | -15.816<br>(22.799) |
|  | 2. In-person=1 hybrid=0 | 26.791**<br>(11.299) | 23.111<br>(15.512) | 12.67<br>(14.917) | 22.044<br>(21.657) |
|  | 3. Hybrid=1 online=0 | -1.979<br>(9.098) | 4.448<br>(12.649) | 3.897<br>(13.085) | -17.015<br>(16.629) |
| 3. PSM | 1. In-person=1 online=0 | 10.704<br>(10.877) | 18.588<br>(14.882) | 4.863<br>(16.026) | -2.444<br>(26.242) |
|  | 2. In-person=1 hybrid=0 | 25.247**<br>(12.406) | 9.418<br>(19.31) | -4.274<br>(16.85) | 7.128<br>(21.691) |
|  | 3. Hybrid=1 online=0 | -1.216<br>(8.448) | -2.301<br>(11.778) | -1.16<br>(12.788) | -25.527<br>(16.251) |
| 4. NN | 1. In-person=1 online=0 | 9.595<br>(10.205) | 12.683<br>(16.835) | 5.308<br>(16.344) | 3.521<br>(20.662) |
|  | 2. In-person=1 hybrid=0 | 11.968<br>(10.81) | 8.134<br>(15.865) | 12.879<br>(15.744) | 41.827**<br>(21.125) |
|  | 3. Hybrid=1 online=0 | -10.882<br>(7.777) | -4.681<br>(9.122) | -6.098<br>(11.438) | -19.883<br>(14.994) |
| 5. KMDM | 1. In-person=1 online=0 | 20.185**<br>(8.953) | 29.215**<br>(11.474) | 39.461***<br>(12.565) | 38.404*<br>(19.662) |
|  | 2. In-person=1 hybrid=0 | 24.329**<br>(10.325) | 26.296*<br>(13.466) | 24.879*<br>(14.868) | 31.853<br>(20.131) |
|  | 3. Hybrid=1 online=0 | 0.727<br>(7.38) | 4.734<br>(9.833) | 15.209<br>(10.742) | 5.31<br>(14.044) |

*Note:* Robust standard errors in parentheses. \*\*\* $p < 0.01$ , \*\* $p < 0.05$ , \* $p < 0.1$ . N=1,076. Model 1 has no control variable. Models 2-5 have the same control variables. Control variables are listed in Table 5 and matching performance is reported in the appendix (Figure A1). State fixed effects were also used but not reported. Biv.=Bivariate, OLS=Ordinary Least Squares, Mult.=Multiple, PSM=Propensity Score Matching, NN=Nearest Neighbor, KMDM=Kernel Multivariate Distance Matching.

Table 3 suggests that the association of in-person teaching with COVID-19 cases, as opposed to online, is positive and significant. For example, the bivariate model shows that cases increase by 18.29 percentage points in the first 15 days after reopening compared to the 15 pre-semester days. However, all other treatments and later periods are not statistically significant in the bivariate model. The gap between the effects of in-person and online stays significant when we control for other covariates, using multiple OLS, PSM, NN, and KMDM respectively; but becomes insignificant for multiple OLS and PSM as we move beyond 30 days. A significant effect in later periods is important from a policy perspective because the difference in cases in the first 15 days can be influenced by mandated testing for in-person teaching mode. Matching with propensity scores does not offer a significant advantage over multiple OLS regression. Matching with NN somewhat corroborates the result and indicates a 48 percentage points increase in two months after reopening. The gap between in-person and hybrid is similar (48 percentage points) by the first two months of reopening, as indicated by the NN model (Table 3).

However, PSM collapses all data dimensions into one, whereas NN is vulnerable to the availability of closest neighbors on all dimensions. A more appropriate and robust method for our analysis is KMDM because it separately performs matching on each dimension. Estimates from NN in Table 3 suggest a statistical difference in COVID-19 cases between in-person and online groups for all periods. For instance, a college reopened with an in-person teaching mode may have 54 percentage points more cases in the county than the pre-opening 15 days in the first month, compared to a similar college with an online teaching mode. The figure becomes 81 percentage points after the first two months.

Interestingly, KMDM results show that the hybrid teaching mode has a positive significant difference from online in later periods. Although the gap between hybrid and online is insignificant in the first month, colleges with hybrid modes have 53 percentage points increase in cases as opposed to colleges with online modes by the end of the second month. The gap between in-person and hybrid remains statistically insignificant. That is, the hybrid mode follows the pattern of in-person after 30 days. To summarize, colleges that chose in-person are associated with a greater increase in COVID-19 cases in their respective counties, compared to the colleges that chose online. A hybrid instructional mode may create a difference from in-person at the beginning, but may not sustain for longer periods, possibly because the spread from in-person components overwhelms the gain from online components due to the exponential nature of the disease spread.

Table 4 presents estimation results for COVID-19 deaths. Relatively simpler models (1-3) show some positive significant differences between in-person and hybrid within the first month. It is possible that colleges with high pre-semester cases influenced the death rates within 30 days when control variables were not properly matched. However, as a more robust method, KMDM captures a statistically significant difference between in-person and online modes, which grows over time at a decreasing rate. For example, teaching in-person as opposed to online is associated with 29 percentage points more deaths in the first 30 days after reopening compared to the 15 pre-semester days, and 38 percentage points more deaths in the first 60 days after reopening. Similar to the results we found for COVID cases, hybrid modes create a statistically significant difference in death rates from in-person for the first 45 days, but the difference gradually dissolves beyond that period.

In short, the magnitude of the average treatment effects increases at a decreasing rate as we go from 0-15 days to 45-60 days in both Tables 3 and 4. This occurs because college reopening increases average cases and deaths regardless of teaching modality. Although online mode raises the rates slower than in-person, the gap starts to shrink after 45 days, possibly because college students are likely to socialize on campus outside the classroom even if the classes are online.

The dataset further allows us to explore another aspect of teaching modality during a pandemic. We use logistic regressions to find the predictors of a teaching modality. The results in Table 5 show that colleges with greater enrollment are less likely to choose in-person than online (column 1), and in-person than hybrid (column 3), holding other variables constant. A percent increase in endowment per student decreases the log odds ratio of choosing in-person over online by 0.616, and that of choosing hybrid over online by 0.476, given the other predictors. More population in the county might have made colleges choose remote teaching components. For example, colleges in counties with a percent more population are associated with 0.812 lower log odds of choosing in-person modes over online. The negative association of in-person mode with enrollment and population variable implies that the colleges might have considered remote teaching elements where campuses are more likely to be crowded.

**Table 5.** Predictors of teaching modalities (Logistic regression coefficients)

| Variables | (1)<br>In-person=1<br>online=0 | (2)<br>Hybrid=1<br>online=0 | (3)<br>In-person=1<br>hybrid=0 |
| --- | --- | --- | --- |
| % stayed home | -0.0289<br>(0.0514) | -0.00890<br>(0.0536) | 0.0122<br>(0.0380) |
| Log enrollment | -0.616***<br>(0.192) | -0.177<br>(0.192) | -0.476***<br>(0.160) |
| Log cost of attendance | 0.568<br>(0.711) | 0.378<br>(0.756) | 0.400<br>(0.696) |
| Log endowment per student | -0.436***<br>(0.152) | -0.388***<br>(0.147) | -0.108<br>(0.103) |
| Log student-faculty ratio | -0.820*<br>(0.444) | -1.125**<br>(0.465) | 0.393<br>(0.385) |
| Public=1, 0 otherwise | -0.895<br>(0.666) | -1.044<br>(0.703) | 0.0880<br>(0.625) |
| Log total population | -0.812***<br>(0.289) | -0.392<br>(0.287) | -0.155<br>(0.239) |
| Black population (%) | 0.00524<br>(0.0203) | 0.00325<br>(0.0204) | 0.0184<br>(0.0170) |
| Male population (%) | -0.103<br>(0.103) | -0.0116<br>(0.116) | -0.0679<br>(0.0870) |
| Log household income | 1.338*<br>(0.753) | 1.018<br>(0.771) | 0.133<br>(0.661) |
| Republican votes (%) | 0.0462***<br>(0.0177) | 0.0156<br>(0.0178) | 0.0346**<br>(0.0154) |
| Mask ordinance=1, 0 otherwise | 1.731<br>(1.126) | -1.075<br>(1.337) | -0.141<br>(1.254) |
| Rural campus=1, 0 otherwise | -0.761<br>(0.478) | -0.278<br>(0.516) | -0.186<br>(0.372) |
| Log cases in spring 2020 | 0.334<br>(0.287) | 0.224<br>(0.272) | 0.00667<br>(0.190) |
| Log deaths in spring 2020 | -0.0715<br>(0.233) | -0.0367<br>(0.216) | -0.121<br>(0.164) |
| Temperature in reopening month | 0.0108<br>(0.0562) | -0.0684<br>(0.0557) | 0.0940<br>(0.0575) |
| County college count | 0.0213<br>(0.0618) | -0.0218<br>(0.0356) | 0.0430<br>(0.0536) |
| Constant | 1.868<br>(11.64) | 1.836<br>(14.27) | -6.738<br>(10.78) |
| Observations (colleges) | 406+386=792 | 284+406=690 | 386+284=670 |
| Accuracy | 78.50% | 70.45% | 69.19% |
| Wald $\chi^2$ | 149.68*** | 75.28** | 78.33** |
| Log pseudolikelihood | -224.37 | -243.49 | -299.97 |
| Pseudo R-squared | 0.334 | 0.168 | 0.152 |

*Note:* Standard errors in parentheses. \*\*\* $p < 0.01$ , \*\* $p < 0.05$ , \* $p < 0.1$ . $N = 1,076$ . The third category is omitted from the sample for each pairwise comparison. For example, 406 online + 386 in-person colleges make 792 colleges in column (1), 284 hybrid and 406 online make 690 colleges in column (2), and so on. State fixed effects were added but not reported. We use log forms of some predictors to reduce the influence of extreme values on coefficient estimates. Accuracy shows the percentage of teaching modalities correctly identified by the model. The pseudo R-squared is higher (0.33) for the in-person versus online prediction (column 1) as opposed to hybrid versus online or in-person versus hybrid. This occurs because hybrid teaching modes are widely heterogeneous across colleges and harder to measure and categorize. Nevertheless, the main interest of this study is to analyze the difference between full in-person and full online, which also gives us directions on their mixture, i.e. hybrid modes. Moreover, pseudo R-squared cannot be interpreted as the goodness of fit because, unlike the R-squared from OLS, it shows the improvement in model likelihood over a null model (e.g., Heinzl et al. 2005; Hu et al. 2006; Hammert et al. 2018). Table 5 also presents the accuracy of the logistic models, suggesting that the teaching modalities of about seven to eight colleges out of 10 colleges were accurately predicted by our model.

Colleges with bigger class sizes, measured by the student-faculty ratio, were less likely to choose in-person teaching modes. The log odds ratio of choosing an in-person mode over online decreases by 0.82 as the student-faculty ratio increases by one percent, whereas that of hybrid decreases by 1.125 over online for the same increase in student-faculty ratio. Table 5 also shows that greater Republican votes in the 2016 election have a positive association with choosing in-person mode over online or hybrid. As discussed in the data section, political views in a county might have played a role in the selection of teaching modality. Interestingly, the real household income in the county has a positive relationship with in-person teaching mode as compared to online. Since college-level financial variables are already controlled for, a possible explanation for this is that wealthier counties preferred in-person over online to reduce the stress and to ensure the quality of education (e.g., Lazarevic and Bentz 2021; Kofoed et al. 2021; Orlov et al. 2021).

The above results are based on the full model that was constructed based on the literature. Some control variables can be correlated, which reduces the precision of estimated coefficients in the logistic model. Figure A2 in the appendix presents the correlation heatmap of control variables. Not surprisingly, total population, COVID-19 cases and deaths in the spring semester are highly correlated (0.90). Since total population is a critical predictor of disease spread, and it also appeared significant in the selection of teaching modes (Table 5), we kept total population in the model and dropped spring cases and deaths variables, and repeated the above analysis for robustness. The results are presented in the appendix Tables A1-A3. Treatment effect estimates in Table A1 and A2 are almost the same and corroborate our previous observations from the KMDM model. The noticeable difference in the logistic model results (Table A3) is that rural campus variable becomes statistically significant; i.e., colleges in rural areas are less likely to choose in-person over online. This can be attributed to the colleges in urban areas that decided to reopen with online modes due to high COVID cases or deaths in the spring semester. Thus, dropping spring outcomes made the rural versus urban variable significant.

## 5 Concluding remarks and implications

What do these results imply? Assume two similar counties with two comparable colleges, and each county reported 100 COVID cases before the fall 2020 semester. Further assume that the first county has its college reopened online, and has 10 more COVID cases in two months. If the college in the second county reopened in person, then its county may have 18 more cases in two months, ceteris paribus. Again, assume 100 COVID-related deaths before the fall 2020 semester in each of these two counties. If the first county observes three deaths in two months, the latter county observes four deaths. Moreover, if the second college reopens with a hybrid mode, then the number of cases and deaths respectively would be 15 and four in two months, because hybrid modes make a difference from in-person only at the beginning, but reaches up to the level of in-person within two months.

We found a statistically significant association between teaching modalities and COVID outcomes, which is consistent with several studies that find a negative impact of remote teaching or school closures on COVID outcomes (e.g., Auger et al. 2020; Pan et al. 2020; Andersen et al. 2020; Chernozhukov et al. 2021), but our result contradicts the studies that find little or no negative impact (e.g., UNICEF 2020; Herby et al. 2022). However, these studies analyze teaching modalities or closures of schools, and not colleges where students are more likely to be affected due to their age and the frequency of socialization (e.g., Wrighton and Lawrence 2020; Goodman et al. 2021). The only exception is Andersen et al. (2020) who analyzed college reopenings. Their preliminary findings suggest that college reopenings increase COVID-19 cases by 1.7 daily cases per 100,000 residents in the first two weeks. Assuming there was no case pre-semester, the exponential growth of 1.7 cases per day for 60 days generates a large number. However, we control for many college and county-level factors that Andersen et al. (2020) did not include in their (preliminary) analysis, which might have contributed to the difference.

Our findings have several implications for higher education and health policy that simultaneously apply to agricultural economics and agribusiness departments and/or land grant universities. First, results from logistic regression suggest that colleges with bigger class sizes, having more populated campuses, or located in more populated areas chose remote teaching components, i.e., preferred online or hybrid to in-person modes. Although the rural campus variable is not a significant predictor in our results, many land-grant universities with urban or semi-urban campuses fall under this category. On the contrary, colleges with smaller endowments per student are less likely to choose online over in-person teaching modes, and hybrid over online modes, after controlling for other factors. Thus, relatively weaker positioned with heavy dependence on traditional on-campus tuition and auxiliary revenues would likely tend to return to the classroom with a quicker frequency out of financial exigency. Moreover, the average treatment effects show adopting an in-person mode instead of an online mode is associated with more COVID-19 cases. Therefore, colleges with small endowments need special policy attention to combat a disease-induced crisis.

Second, our results indicate that wealthier communities are more likely to have in-person teaching modalities, then less wealthy communities must have resources for online teaching. However, many studies discuss the added stress and the lack of social interactions in online instruction modes and hence poor student performance (e.g., Picault 2021; Lazarevic and Bentz 2021; Kofoed et al. 2021; Orlov et al. 2021). Thus, communities with low real income are in greater need of teaching resources on making online courses more interactive and performance-oriented.

Third, the analysis also suggests that campus reopening has a positive relationship with COVID-19 cases across all teaching modes, and both new cases and new deaths tend to increase for colleges teaching in-person compared to colleges teaching online. Since the treatment effects ideally find counterfactuals, one can point out that the expected risk of spreading a communicable disease can be partially mitigated with an initiative from the colleges by increasing distance education elements in classes. For hybrid modes, it is important to follow the containment policies at both the college and community level, as we found that the in-person components of hybrid modes may influence the disease spread and undermine the expected benefits from remote instruction.

Our study is observational and not based on a randomized placebo-controlled trial, hence should be interpreted with great caution. However, it still finds significant patterns in the data, offering important insights about choosing a teaching modality during a disease-induced pandemic, given the college and county-level features. Future research can look into the short-run and long-run effects of different teaching modalities on student learning outcomes.

## Data Availability

All data used are publicly available and do not include any individual level information.

## Acknowledgements

Authors would like to thank the Editor and Reviewers, and the participants of the American Economic Association annual meeting 2022 for their helpful comments. Authors have no conflict of interests. No human subject was involved in this research. No funding was received.

### Appendix

**Figure A1.**
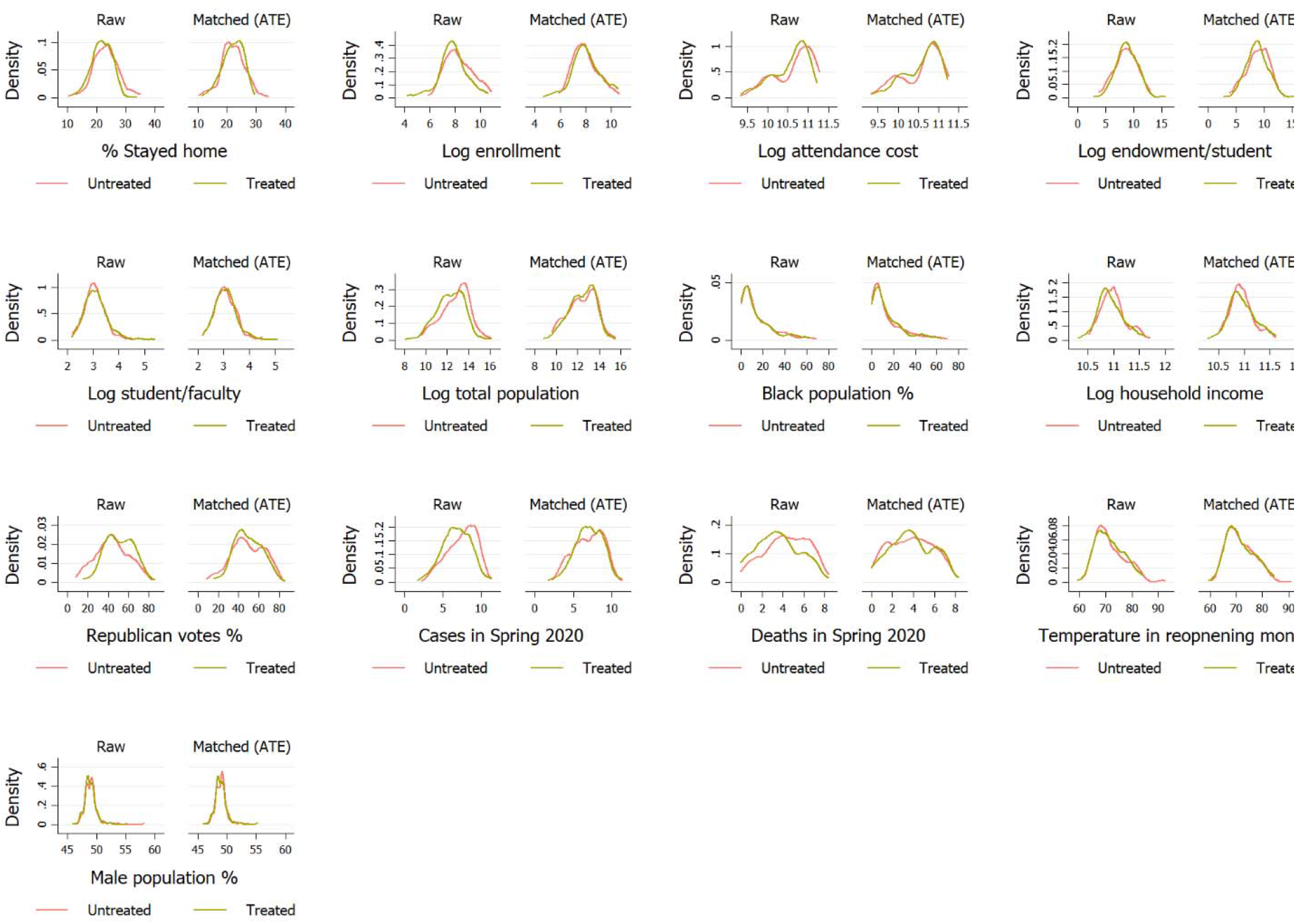
Densities of major variables before and after matching. *Note:* Figure shows kernel matching for multiple variables. See Table 1 for variable description and Table 2 for summary statistics.

**Figure A2.**
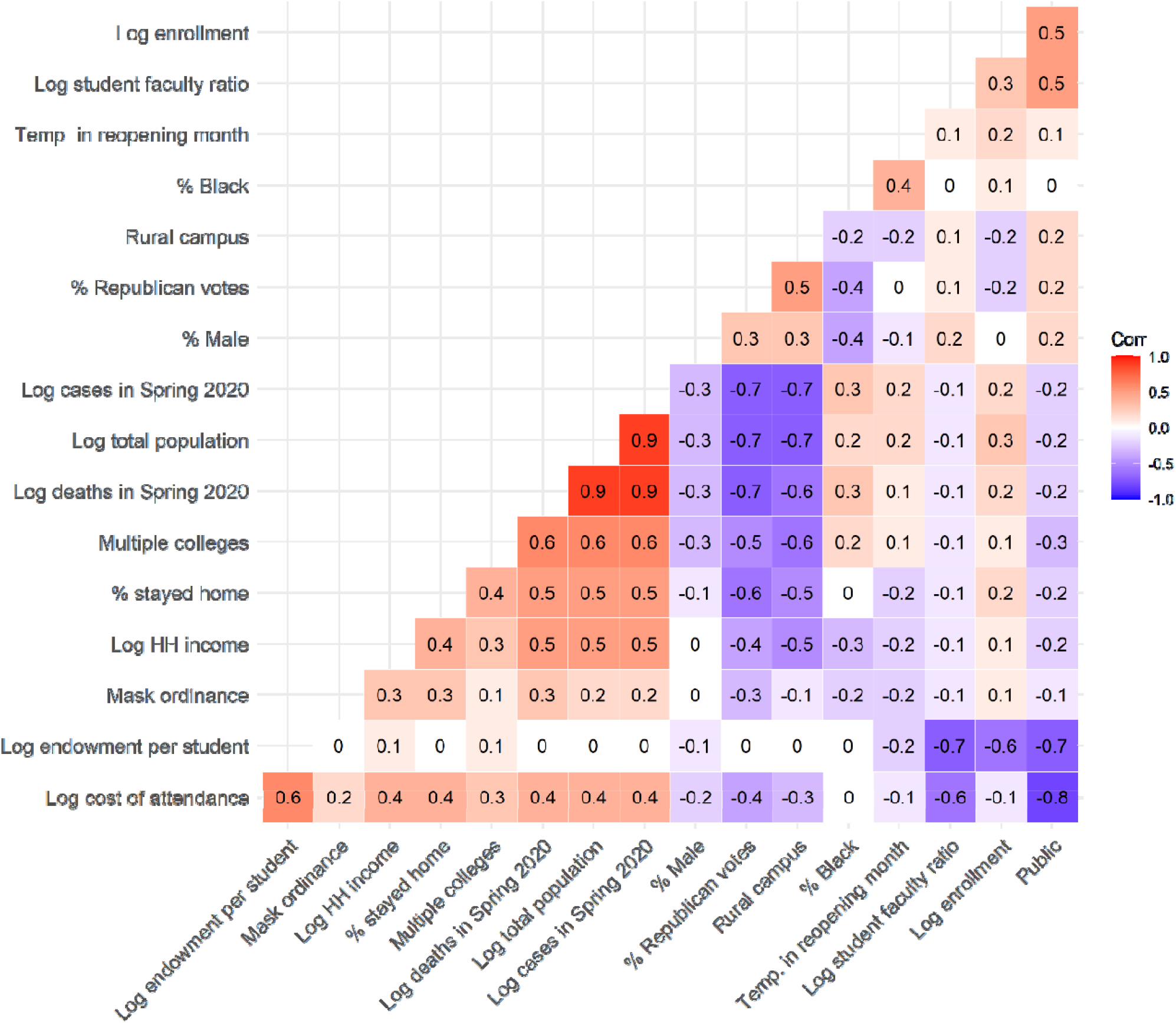
Correlation heatmap of control variables. *Note*: Figure shows Pearson correlation coefficients, rounded to one digit after the decimal and ordered by the strength of correlation. Some figures are too small and become zero when rounded. Darker shades mean greater association. See Table 1 for variable description and Table 2 for summary statistics.

**Table A1.** Average treatment effects on COVID-19 cases. (Dependent variable: % change in cases compared to cases in two weeks before reopening date)

| Model | Treatment | (1) | (2) | (3) | (4) |
| --- | --- | --- | --- | --- | --- |
|  |  | 0-15 Days | 15-30 Days | 30-45 Days | 45-60 Days |
| 1. Biv. OLS | 1. In-person=1 online=0 | 18.291**<br>(9.259) | 22.265<br>(13.625) | 38.044<br>(24.626) | 24.598<br>(29.549) |
|  | 2. In-person=1 hybrid=0 | 12.768<br>(10.273) | 3.537<br>(20.489) | 27.221<br>(30.055) | 21.753<br>(34.139) |
|  | 3. Hybrid=1 online=0 | 5.552<br>(8.497) | 18.532<br>(16.705) | 22.661<br>(17.007) | 19.474<br>(23.512) |
| 2. Mult. OLS | 1. In-person=1 online=0 | 31.273**<br>(14.957) | 36.068*<br>(20.95) | 54.87<br>(35.865) | 1.804<br>(37.521) |
|  | 2. In-person=1 hybrid=0 | 24.88*<br>(12.798) | 21.364<br>(21.711) | 43.458<br>(34.555) | 4.768<br>(37.635) |
|  | 3. Hybrid=1 online=0 | 3.229<br>(12.012) | 9.693<br>(18.04) | 18.837<br>(16.673) | 36.561<br>(31.07) |
| 3. PSM | 1. In-person=1 online=0 | 41.677***<br>(15.275) | 41.11**<br>(20.893) | 61.454<br>(37.993) | 16.121<br>(45.06) |
|  | 2. In-person=1 hybrid=0 | 27.596**<br>(12.524) | 24.916<br>(22.504) | 44.545<br>(36.494) | 11.999<br>(41.782) |
|  | 3. Hybrid=1 online=0 | 1.906<br>(9.34) | 5.135<br>(14.837) | 16.283<br>(17.925) | 27.897<br>(37.268) |
| 4. NN | 1. In-person=1 online=0 | 27.616**<br>(13.409) | 29.32*<br>(15.895) | 46.174<br>(28.347) | 43.329<br>(31.894) |
|  | 2. In-person=1 hybrid=0 | 23.906**<br>(10.426) | 28.757<br>(19.796) | 63.644<br>(41.519) | 71.883<br>(54.532) |
|  | 3. Hybrid=1 online=0 | 7.947<br>(11.577) | 13.505<br>(18.526) | 13.993<br>(15.835) | 28.416<br>(32.107) |
| 5. KMDM | 1. In-person=1 online=0 | 35.414***<br>(10.769) | 55.546***<br>(15.509) | 80.956***<br>(28.45) | 83.528**<br>(36.497) |
|  | 2. In-person=1 hybrid=0 | 18.504<br>(13.133) | 13.74<br>(23.634) | 38.934<br>(31.916) | 25.924<br>(41.911) |
|  | 3. Hybrid=1 online=0 | 14.418<br>(11.016) | 27.957<br>(19.9) | 33.382**<br>(16.667) | 52.470*<br>(29.037) |

*Note:* Robust standard errors in parentheses. \*\*\* p<0.01, \*\* p<0.05, \* p<0.1. N=1,076. Model 1 has no control variable. Models 2-5 have the same control variables. Control variables are listed in Table 5 and matching performance is reported in the appendix (Figure A1). State fixed effects were also used but not reported. Biv.=Bivariate, OLS=Ordinary Least Squares, Mult.=Multiple, PSM=Propensity Score Matching, NN=Nearest Neighbor, KMDM=Kernel Multivariate Distance Matching.

**Table A2.**
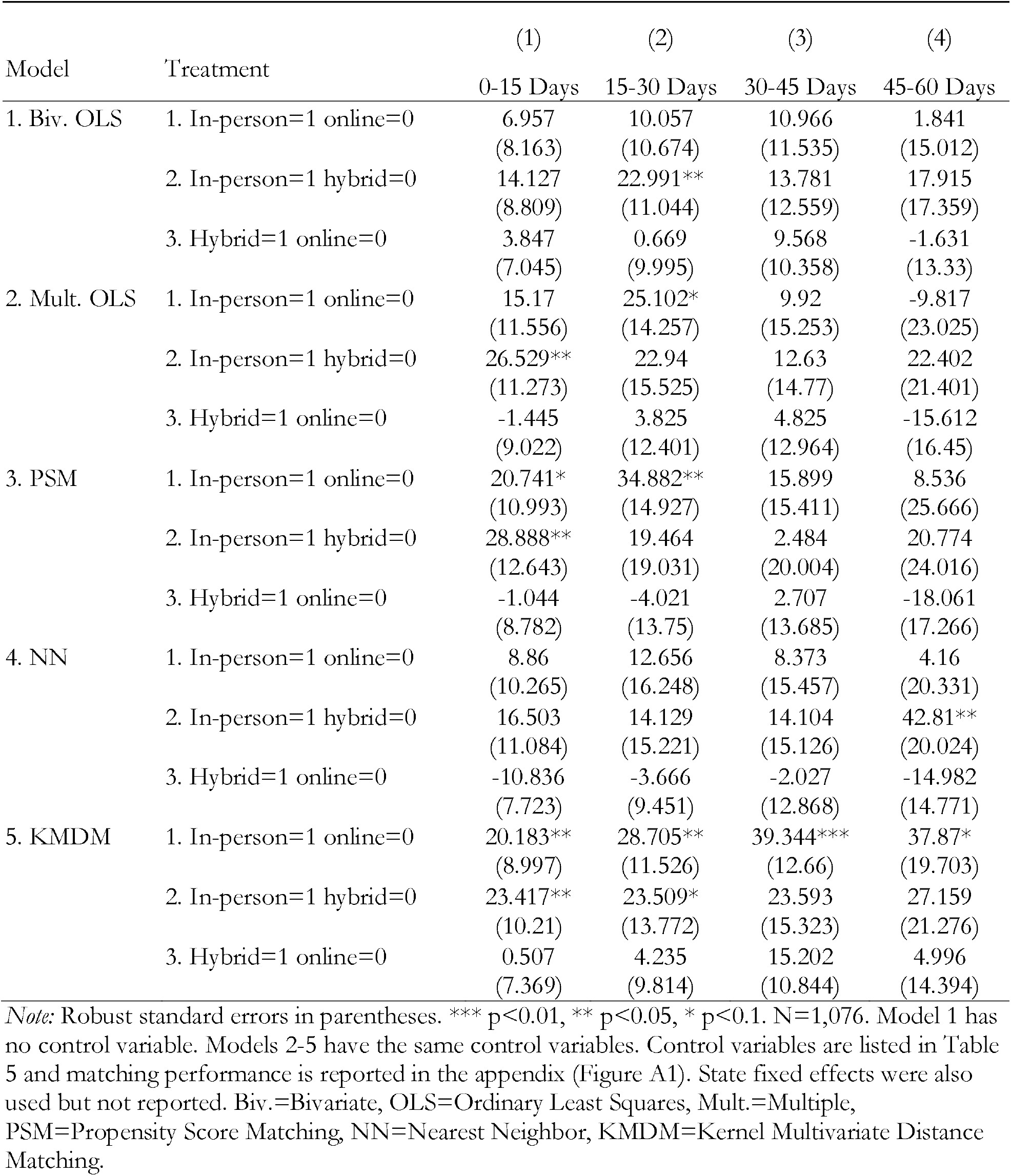
Average treatment effects on COVID-19 deaths. (Dependent variable: % change in deaths compared to deaths in two weeks before reopening date)

| Model | Treatment | (1) | (2) | (3) | (4) |
| --- | --- | --- | --- | --- | --- |
|  |  | 0-15 Days | 15-30 Days | 30-45 Days | 45-60 Days |
| 1. Biv. OLS | 1. In-person=1 online=0 | 6.957<br>(8.163) | 10.057<br>(10.674) | 10.966<br>(11.535) | 1.841<br>(15.012) |
|  | 2. In-person=1 hybrid=0 | 14.127<br>(8.809) | 22.991**<br>(11.044) | 13.781<br>(12.559) | 17.915<br>(17.359) |
|  | 3. Hybrid=1 online=0 | 3.847<br>(7.045) | 0.669<br>(9.995) | 9.568<br>(10.358) | -1.631<br>(13.33) |
| 2. Mult. OLS | 1. In-person=1 online=0 | 15.17<br>(11.556) | 25.102*<br>(14.257) | 9.92<br>(15.253) | -9.817<br>(23.025) |
|  | 2. In-person=1 hybrid=0 | 26.529**<br>(11.273) | 22.94<br>(15.525) | 12.63<br>(14.77) | 22.402<br>(21.401) |
|  | 3. Hybrid=1 online=0 | -1.445<br>(9.022) | 3.825<br>(12.401) | 4.825<br>(12.964) | -15.612<br>(16.45) |
| 3. PSM | 1. In-person=1 online=0 | 20.741*<br>(10.993) | 34.882**<br>(14.927) | 15.899<br>(15.411) | 8.536<br>(25.666) |
|  | 2. In-person=1 hybrid=0 | 28.888**<br>(12.643) | 19.464<br>(19.031) | 2.484<br>(20.004) | 20.774<br>(24.016) |
|  | 3. Hybrid=1 online=0 | -1.044<br>(8.782) | -4.021<br>(13.75) | 2.707<br>(13.685) | -18.061<br>(17.266) |
| 4. NN | 1. In-person=1 online=0 | 8.86<br>(10.265) | 12.656<br>(16.248) | 8.373<br>(15.457) | 4.16<br>(20.331) |
|  | 2. In-person=1 hybrid=0 | 16.503<br>(11.084) | 14.129<br>(15.221) | 14.104<br>(15.126) | 42.81**<br>(20.024) |
|  | 3. Hybrid=1 online=0 | -10.836<br>(7.723) | -3.666<br>(9.451) | -2.027<br>(12.868) | -14.982<br>(14.771) |
| 5. KMDM | 1. In-person=1 online=0 | 20.183**<br>(8.997) | 28.705**<br>(11.526) | 39.344***<br>(12.66) | 37.87*<br>(19.703) |
|  | 2. In-person=1 hybrid=0 | 23.417**<br>(10.21) | 23.509*<br>(13.772) | 23.593<br>(15.323) | 27.159<br>(21.276) |
|  | 3. Hybrid=1 online=0 | 0.507<br>(7.369) | 4.235<br>(9.814) | 15.202<br>(10.844) | 4.996<br>(14.394) |

*Note:* Robust standard errors in parentheses. \*\*\* $p < 0.01$ , \*\* $p < 0.05$ , \* $p < 0.1$ . N=1,076. Model 1 has no control variable. Models 2-5 have the same control variables. Control variables are listed in Table 5 and matching performance is reported in the appendix (Figure A1). State fixed effects were also used but not reported. Biv.=Bivariate, OLS=Ordinary Least Squares, Mult.=Multiple, PSM=Propensity Score Matching, NN=Nearest Neighbor, KMDM=Kernel Multivariate Distance Matching.

**Table A3.** Predictors of teaching modalities (Logistic regression coefficients)

| Variables | (1)<br>In-person=1<br>online=0 | (2)<br>Hybrid=1<br>online=0 | (3)<br>In-person=1<br>hybrid=0 |
| --- | --- | --- | --- |
| % stayed home | -0.035<br>(0.047) | -0.008<br>(0.053) | 0.009<br>(0.038) |
| Log enrollment | -0.612***<br>(0.186) | -0.192<br>(0.190) | -0.459***<br>(0.158) |
| Log cost of attendance | 0.516<br>(0.715) | 0.386<br>(0.752) | 0.413<br>(0.692) |
| Log endowment per student | -0.420***<br>(0.145) | -0.400***<br>(0.144) | -0.099<br>(0.103) |
| Log student-faculty ratio | -0.758*<br>(0.438) | -1.130**<br>(0.462) | 0.401<br>(0.382) |
| Public=1, 0 otherwise | -0.894<br>(0.661) | -1.067<br>(0.699) | 0.092<br>(0.623) |
| Log total population | -0.522**<br>(0.227) | -0.183<br>(0.196) | -0.283<br>(0.176) |
| Black population (%) | 0.009<br>(0.0197) | 0.006<br>(0.019) | 0.016<br>(0.016) |
| Male population (%) | -0.104<br>(0.0997) | -0.004<br>(0.116) | -0.069<br>(0.087) |
| Log household income | 1.565**<br>(0.749) | 1.195<br>(0.744) | 0.110<br>(0.655) |
| Republican votes (%) | 0.0434**<br>(0.018) | 0.014<br>(0.017) | 0.0348**<br>(0.015) |
| Mask ordinance=1, 0 otherwise | 1.680<br>(1.099) | -0.933<br>(1.332) | -0.232<br>(1.240) |
| Rural campus=1, 0 otherwise | -0.794*<br>(0.465) | -0.280<br>(0.507) | -0.190<br>(0.371) |
| Temperature in reopening month | 0.037<br>(0.056) | -0.058<br>(0.052) | 0.082<br>(0.055) |
| County college count | 0.027<br>(0.061) | -0.018<br>(0.035) | 0.039<br>(0.053) |
| Constant | -3.388<br>(11.410) | -2.391<br>(12.910) | -4.542<br>(10.280) |
| Observations (colleges) | 406+386=792 | 284+406=690 | 386+284=670 |
| Accuracy | 78.38% | 70.21% | 67.89% |
| Wald $\chi^2$ | 140.87*** | 74.16** | 77.57** |
| Log pseudolikelihood | -225.72 | -244.05 | -300.49 |
| Pseudo R-squared | 0.332 | 0.166 | 0.152 |

*Note:* Standard errors in parentheses. \*\*\* $p < 0.01$ , \*\* $p < 0.05$ , \* $p < 0.1$ . $N = 1,076$ . The third category is omitted from the sample for each pairwise comparison. For example, 406 online + 386 in-person colleges make 792 colleges in column (1). State fixed effects were added but not reported. We use log forms of some predictors to reduce the influence of extreme values on coefficient estimates. Accuracy shows the percentage of teaching modalities correctly identified by the model.

## Footnotes

1 The motivation of addressing the estimation issues is to try our best to make sure that the treatment assignment was, at least statistically, “equally likely” and the estimates are not biased. A randomized placebo-controlled trail would have generated a more accurate estimate. We discuss this limitation in the concluding section.

2 Cellphone locations are accurate up to 30 meters on average, and are recorded in every 10 minutes for 24/7. The percentage of people went outside home means people who went farther than 30 meters from their night-staying location at any point in 24 hours for longer than 10 minutes. The implicit assumption we make here is at least college students have access to a cellphone device. This is not unrealistic because Pew Research Center (2021) reports that 100% of Americans aged 18-29 own a cellphone of some kind. Another assumption is to define fixed night-staying location as home.

3 A list of colleges and their webpages are publicly available in school_info.txt on https://github.com.

